# Development and Validation of a Type 1 Diabetes Multi-Ancestry Polygenic Score

**DOI:** 10.1101/2025.06.20.25329522

**Authors:** Aaron J. Deutsch, Andrew S. Bell, Dominika A. Michalek, Adam B. Burkholder, Stella Nam, Raymond J. Kreienkamp, Seth A. Sharp, Alicia Huerta-Chagoya, Ravi Mandla, Ruth Nanjala, Yang Luo, Richard A. Oram, Jose C. Florez, Suna Onengut-Gumuscu, Stephen S. Rich, Alison A. Motsinger-Reif, Alisa K. Manning, Josep M. Mercader, Miriam S. Udler

## Abstract

**Objective:** Polygenic scores strongly predict type 1 diabetes risk, but most scores were developed in European-ancestry populations. In this study, we developed a multi-ancestry polygenic score to accurately predict type 1 diabetes risk across diverse populations.

**Research Design and Methods:** We used recent multi-ancestry genome-wide association studies to create a type 1 diabetes multi-ancestry polygenic score (T1D MAPS). We trained the score in the Mass General Brigham (MGB) Biobank (372 individuals with type 1 diabetes) and tested the score in the All of Us program (86 individuals with type 1 diabetes). We evaluated the area under the receiver operating characteristic curve (AUC), and we compared the AUC to two published single-ancestry scores: T1D GRS2_EUR_ and T1D GRS_AFR_. We also developed an updated score (T1D MAPS2) that combines T1D GRS2_EUR_ and T1D MAPS.

**Results:** Among individuals with non-European ancestry, the AUC of T1D MAPS was 0.90, significantly higher than T1D GRS2_EUR_ (0.82, *P* = 0.04) and T1D GRS_AFR_ (0.82, *P* = 0.007). Among individuals with European ancestry, the AUC of T1D MAPS was slightly lower than T1D GRS2_EUR_ (0.89 vs. 0.91, *P* = 0.02). However, T1D MAPS2 performed equivalently to T1D GRS2_EUR_ in European ancestry (0.91 vs. 0.91, *P* = 0.45) while still performing better in non-European ancestry (0.90 vs. 0.82, *P* = 0.04).

**Conclusions:** A novel polygenic score improves type 1 diabetes risk prediction in non-European ancestry while maintaining high predictive power in European ancestry. These findings advance the accuracy of type 1 diabetes genetic risk prediction across diverse populations.

**Article Highlights:**

- Why did we undertake this study? Type 1 diabetes polygenic scores are highly predictive of disease risk, but their performance varies based on genetic ancestry.
- What is the specific question(s) we wanted to answer? Can we develop a polygenic score that accurately predicts type 1 diabetes risk across diverse populations?
- What did we find? Our novel polygenic score performs similarly to existing scores in European populations, and it demonstrates superior performance in non-European populations.
- What are the implications of our findings? This polygenic score will improve prediction of type 1 diabetes risk in genetically diverse populations.

---

Type 1 diabetes is a complex disease with multiple genetic and environmental risk factors. Strategies to identify individuals at high risk of type 1 diabetes can promote earlier detection, reducing diabetes-related morbidity and identifying potential candidates for disease-modifying monoclonal antibody therapy^1^. Furthermore, the diagnosis of type 1 diabetes may be overlooked among individuals with known diabetes; while type 1 diabetes is classically considered a disease of childhood onset, the disease can occur throughout adulthood^2^ and may be misdiagnosed, particularly in individuals with atypical presentations^3–5^.

Polygenic scores – which integrate the effects of multiple variants across the genome – offer a powerful approach to determine disease risk. Current polygenic scores display an outstanding ability to distinguish between individuals with and without type 1 diabetes, with an area under the receiver operating curve (AUC) of 0.9 or greater in individuals with European genetic ancestry^6^. However, because existing polygenic scores were primarily developed in European populations, their accuracy may decrease when applied to other ancestry groups^7,8^ or to genetically admixed populations^9–11^. Furthermore, the distribution of polygenic scores may differ across ancestry groups, creating a need for ancestry-specific score thresholds^12^. Alternatively, novel polygenic scores can incorporate ancestry-specific risk variants to improve disease prediction in specific populations^13,14^. However, this approach requires investigators to select the optimal ancestry-matched polygenic score, which can be challenging, particularly for individuals from admixed genetic backgrounds.

Concerns about the transferability of polygenic scores have motivated calls to increase population diversity in genetic studies^15^ and develop methods to capture disease risk across diverse populations^16–19^. Multi-ancestry risk prediction is particularly challenging in type 1 diabetes because disease risk is strongly influenced by the human leukocyte antigen (HLA) genes, which exhibit substantial allelic variation across global populations^20^.

In this study, we leveraged recent diverse genetic studies to create a novel risk score, Type 1 Diabetes Multi-Ancestry Polygenic Score (T1D MAPS). Compared to existing polygenic scores, T1D MAPS demonstrates equivalent predictive power for individuals with European ancestry and improved predictive power for individuals with non-European ancestry, including African and Admixed American ancestry. Notably, we observed significant results despite a modest type 1 diabetes sample size in our training and testing cohorts. Furthermore, the distribution of T1D MAPS is similar across populations, allowing for a single universal score threshold to identify high-risk individuals. These results advance our understanding of type 1 diabetes risk and will help to ensure optimal care for all individuals with type 1 diabetes.

## Research Design and Methods

### Construction of type 1 diabetes polygenic score

We focused separately on the major histocompatibility complex (MHC), which includes the HLA genes that strongly determine type 1 diabetes risk, as well as variants outside the MHC (Fig. 1A). We used publicly available summary statistics from a recent multi-ancestry genome-wide association study (GWAS) of type 1 diabetes (discovery cohort 1), which included over 4,000 individuals with type 1 diabetes comprising three genetic ancestry groups: African (AFR), Admixed American (AMR), and European (EUR)^21^. To analyze genetic variants outside the MHC region, we used a recent EUR-ancestry GWAS that included approximately 19,000 individuals with type 1 diabetes (discovery cohort 2)^22^. We combined the HLA and non-HLA scores to determine the overall score.

**Figure 1.**
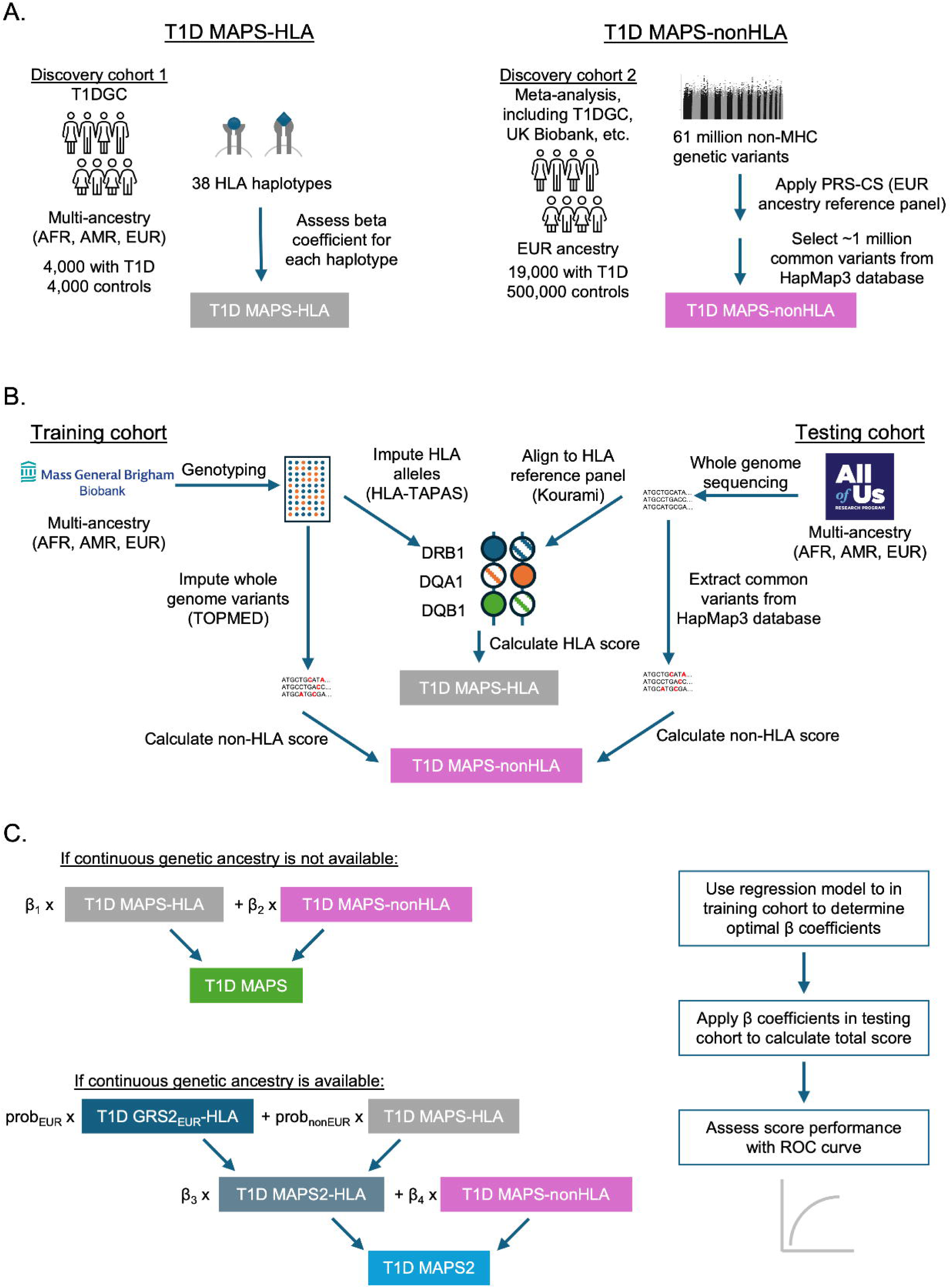
Overview of experimental design. (A) Description of discovery cohorts used to select weights for T1D MAPS-HLA and T1D MAPS-nonHLA. (B) Approach used to calculate T1D MAPS-HLA and T1D MAPS-nonHLA in training and testing cohorts. (C) Approach used to combine T1D MAPS-HLA and T1D MAPS-nonHLA into overall T1D MAPS or T1D MAPS2 score. T1DGC, Type 1 Diabetes Genetics Consortium.

### Biobank cohorts

To train the polygenic score, we used the Mass General Brigham (MGB) Biobank, a repository linked to electronic medical records at the Mass General Brigham hospital system in Boston, Massachusetts^23^ (Fig. 1B). Analysis of MGB Biobank was approved by the MGB IRB (study protocol 2016P001018). Individuals were genotyped using the Illumina Multi-Ethnic Genotyping Array or the Illumina Infinium Global Screening Array^24^. Imputation was performed using the Trans-Omics for Precision Medicine (TOPMED) reference panel^25^. Data from MGB Biobank were current as of October 2022.

To validate the score, we used the All of Us research program, a longitudinal cohort study across the United States with detailed survey data and health information^26^ (Fig. 1B). Analysis of the All of Us cohort was approved by an institutional Data Use and Registration Agreement between MGB and the All of Us Research Program (study protocol 2020P002213). Individuals underwent short-read whole genome sequencing^27^. We performed analyses with the All of Us Controlled Tier Dataset v7 release. Both MGB Biobank and All of Us were independent of the discovery cohorts used to identify type 1 diabetes genetic associations.

### Phenotype definitions

#### MGB Biobank

Type 1 diabetes was defined based on manual review of medical records by a trained medical reviewer, as described previously^28^. Briefly, individuals needed to meet all of the following criteria: type 1 diabetes diagnosis confirmed by an endocrinologist or primary care physician; current use of basal/bolus insulin regimen or insulin pump; and no secondary cause of diabetes listed in the medical record, such as pancreatitis or glucocorticoid use. Type 2 diabetes was defined using a phenotype algorithm developed by MGB Biobank, with a set positive predictive value of 0.95.

#### All of Us

Individual-level clinical notes were not available in All of Us. Therefore, type 1 diabetes was defined based on structured data available in the medical record, as described previously^29^. Briefly, individuals needed to meet all of the following criteria: presence of type 1 diabetes diagnosis code before age 30, presence of insulin prescription, and absence of prescription for non-insulin glucose-lowering agents. As a secondary analysis, we tested a type 1 diabetes phenotype algorithm developed by the Electronic Medical Records and Genomics (eMERGE) consortium (“T1D-EHR”)^30^. We also implemented a separate algorithm adapted from the eMERGE consortium to define type 2 diabetes (“T2D-EHR+”)^30^.

### Classification of genetic ancestry

We performed principal component analysis in MGB Biobank and projected the principal components onto a diverse reference panel from the Human Genome Diversity Panel (HGDP) and the 1000 Genomes Project^31^. We used a random forest classifier to assign participants to one of six continental ancestry groups: African (AFR),

Latino/admixed American (AMR), East Asian (EAS), Middle Eastern (MID), European (EUR), or South Asian (SAS). This approach generated a continuous probability for each ancestry group. For categorical assignments, individuals were assigned to the ancestry group with the highest probability. The All of Us Research Program used a similar approach to determine genetic ancestry, and results were made available to all investigators using the Researcher Workbench^27^.

The demographic distribution of each dataset is provided in Supplementary Table S1. To allow for statistical testing, we only analyzed ancestry groups that included at least five individuals with type 1 diabetes. This resulted in the following ancestry groups: AFR, AMR, and EUR. In accordance with the All of Us Data and Statistics Dissemination Policy, no data or aggregate statistics corresponding to fewer than 20 participants are displayed; therefore, all genetic ancestries aside from European were combined into a single category labeled as “non-European.”

### Generation of HLA haplotypes

In MGB Biobank, we used genotype array data to impute classical HLA alleles with HLA-TAPAS^32,33^, using an HLA reference panel that includes 21,546 whole-genome sequences spanning five global populations. We then constructed phased multi-locus HLA class II haplotypes, including *HLA*-*DRB1*, *HLA-DQA1*, and *HLA-DQB1*.

In All of Us, we assembled HLA classical alleles from whole genome sequencing data using Kourami^34^. We then performed phasing by constructing each of four possible diploid genotypes across *HLA*-*DRB1*, *HLA-DQA1*, and *HLA-DQB1*, resulting in eight possible haplotypes. We compared each possible haplotype to the list of 38 HLA haplotypes included in the discovery cohort^21^. If an individual had more than two possible haplotypes listed in the discovery cohort, or if more than one possible genotype could be constructed from the available haplotypes, that individual was assigned an HLA score of zero (i.e. disease risk equivalent to the population mean). The total number of haplotypes included for each individual is displayed in Supplementary Table S2.

### Construction of T1D MAPS-HLA

We assigned a score for each of 38 *HLA*-*DRB1-DQA1-DQB1* haplotypes based on the log odds of association with type 1 diabetes in discovery cohort 1^21^ (Supplementary Table S3). If more than one ancestry-specific odds ratio was available, we assigned a single score for each haplotype by taking the log odds from the population with the largest available sample size in the discovery cohort (EUR > AFR > AMR). Any haplotype not found in the discovery cohort was assigned a score of zero.

In addition, we developed a second version of the HLA score, T1D MAPS2-HLA, by calculating an average of T1D MAPS-HLA and T1D GRS2_EUR_, weighted by the percent predicted EUR ancestry:

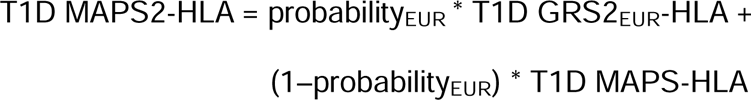

### Construction of T1D MAPS-nonHLA

For the non-HLA score, we used GWAS summary statistics from discovery cohort 2^22^. We excluded variants located in the MHC region, defined as chr6:28,000,000-34,000,000 in GRCh37 coordinates. We implemented PRS-CS^35^, which applies a continuous shrinkage parameter (phi) to generate a global extended polygenic score, using a EUR-ancestry reference panel from 1000 Genomes. We selected approximately 1 million genetic variants found in reference panels from the HapMap3 consortium. Using MGB Biobank as a training cohort, we tested the following phi values: 10^-2^, 10^-3^, 10^-4^, 10^-5^, 10^-6^.

As a secondary analysis, we implemented PRS-CSx^36^, which extends PRS-CS by integrating GWAS data across multiple ancestries. We used AFR and AMR summary statistics from discovery cohort 1^21^ as well as EUR summary statistics from discovery cohort 2^22^. Likewise, we used AFR-, AMR-, and EUR-ancestry reference panels from 1000 Genomes. Then, using MGB Biobank as a training cohort, we constructed a logistic regression model, and we applied the resulting beta coefficients to calculate the non-HLA score as a weighted average of the three ancestry-specific scores.

### Construction of overall T1D MAPS score

For the overall T1D MAPS score, we used MGB Biobank as a training cohort (Fig. 1C). We ran a logistic regression model and calculated the overall score as a weighted average of the HLA score and the non-HLA score:

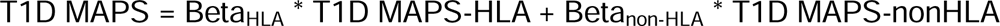

Then, we applied the same beta coefficients to generate the overall T1D MAPS score in All of Us. As a sensitivity analysis, we calculated the T1D MAPS score with and without four principal components of genetic ancestry in the regression model. We used a similar approach to calculate T1D MAPS2 as a weighted average of T1D MAPS2-HLA and T1D MAPS-nonHLA.

### Evaluation of polygenic score performance

We evaluated each polygenic score with the area under the receiver operating characteristic curve (AUC). As a comparison, we implemented two previously published polygenic scores: T1D GRS2_EUR_, which was developed in EUR ancestry^6^, and T1D GRS_AFR_, which was developed in AFR ancestry^13^. For our primary analysis, we compared the AUC of T1D MAPS to T1D GRS2_EUR_ using the DeLong test. As secondary analyses, we also compared the AUCs for different components of each polygenic score (HLA vs. non-HLA), and we assessed the AUC of each polygenic score in a logistic regression model that included four principal components of genetic ancestry. Because HLA allele frequency is highly correlated with genetic ancestry^20^, we chose to focus our primary analysis on the AUC of each polygenic score on its own, following prior studies^6,13^.

## Results

### Development of HLA score

We trained T1D MAPS in MGB Biobank (Fig. 1B), which contained approximately 64,000 individuals, including 372 individuals with type 1 diabetes (335 with EUR ancestry and 37 with non-EUR ancestry; Supplementary Table S1). First, we imputed HLA haplotypes from genotype array data. We found that the genetic variants used to tag HLA haplotypes in T1D GRS2_EUR_ were highly correlated with imputed haplotypes among individuals with EUR ancestry, but the correlation was weaker in non-EUR ancestry (mean R^2^ 0.90 vs. 0.75, Wilcoxon *P* = 9.1 x 10^-3^; Supplementary Fig. S1A). This supported our hypothesis that a multi-ancestry score may better capture HLA allelic variation among non-EUR populations.

Next, we calculated T1D MAPS-HLA using the beta coefficients from the HLA haplotypes in the discovery cohort. We compared the AUC of T1D MAPS-HLA with the HLA components of T1D GRS2_EUR_ and T1D GRS_AFR_ (Supplementary Fig. S2A). T1D GRS_AFR_-HLA had the lowest AUC (0.80-0.82). Among individuals with non-EUR ancestry, the AUC of T1D MAPS-HLA was not significantly different than T1D GRS2_EUR_-HLA (0.89 vs. 0.84; *P* = 0.08, DeLong test). However, among individuals with EUR ancestry, T1D MAPS-HLA had a marginally lower AUC than T1D GRS2_EUR_-HLA (0.84 vs. 0.86, *P* = 0.04). To address this discrepancy, we developed an updated score, T1D MAPS2-HLA, which comprises a weighted average of T1D MAPS-HLA and T1D GRS2_EUR_-HLA based on an individual’s predicted proportion of EUR genetic ancestry. In EUR ancestry, T1D MAPS2-HLA performed marginally better than T1D GRS2_EUR_-HLA (AUC = 0.862 vs. 0.861, *P* = 0.03). In non-EUR ancestry, there was no significant difference between T1D MAPS2-HLA and T1D GRS2_EUR_-HLA (AUC = 0.89 vs. 0.84, *P* = 0.06).

### Development of non-HLA score

We then evaluated T1D MAPS-nonHLA in MGB Biobank. We applied PRS-CS using multiple values for the continuous shrinkage parameter (phi), and we observed the optimal performance with phi = 10^-5^ (Supplementary Table S4). Next, we attempted to integrate GWAS data across multiple ancestries using using PRS-CSx^36^. We used AFR and AMR summary statistics from discovery cohort 1^21^ and EUR summary statistics from discovery cohort 2^22^. However, we found that the AUC using PRS-CSx was lower compared to the results using PRS-CS (EUR ancestry, 0.68 vs. 0.73, *P* = 9.9 x 10^-4^; non-EUR ancestry, 0.62 vs. 0.66, *P* = 0.29). Therefore, we chose to focus on the non-HLA score produced by PRS-CS.

When we assessed the performance in MGB Biobank, we found that T1D MAPS-nonHLA had a significantly higher AUC than T1D GRS2_EUR_-nonHLA within EUR ancestry (0.73 vs. 0.65, *P* = 1.7 x 10^-6^; Supplementary Fig. S2B). In non-EUR ancestry, the AUC of T1D MAPS-nonHLA was not significantly different than T1D GRS2_EUR_-nonHLA (0.66 vs. 0.56, *P* = 0.18). T1D GRS_AFR_-nonHLA had the lowest AUC (0.54-0.58), likely because this score only includes two genetic variants.

### Development of overall T1D MAPS

To construct the overall polygenic score, we applied a logistic regression model in MGB Biobank to model type 1 diabetes status as a function of T1D MAPS-HLA and T1D MAPS-nonHLA. First, we assessed the AUC of each overall score in MGB Biobank, acknowledging the possibility of overfitting (Fig. 2A). Within EUR ancestry, T1D MAPS and T1D GRS2_EUR_ had equivalent AUC (0.88 vs. 0.88, *P* = 0.90), whereas T1D MAPS2 showed superior performance (0.90 vs. 0.88, *P* = 3.0 x 10^-3^). Among non-EUR ancestry, T1D MAPS and T1D MAPS2 were both superior to T1D GRS2 (0.89 vs. 0.80, *P* = 0.01; 0.90 vs. 0.80, *P* = 7.3 x 10^-3^). Both scores also outperformed T1D GRS_AFR_ in this population (0.89 vs. 0.81, *P* = 1.8 x 10^-3^; 0.90 vs. 0.81, *P* = 8.4 x 10^-4^).

**Figure 2.**
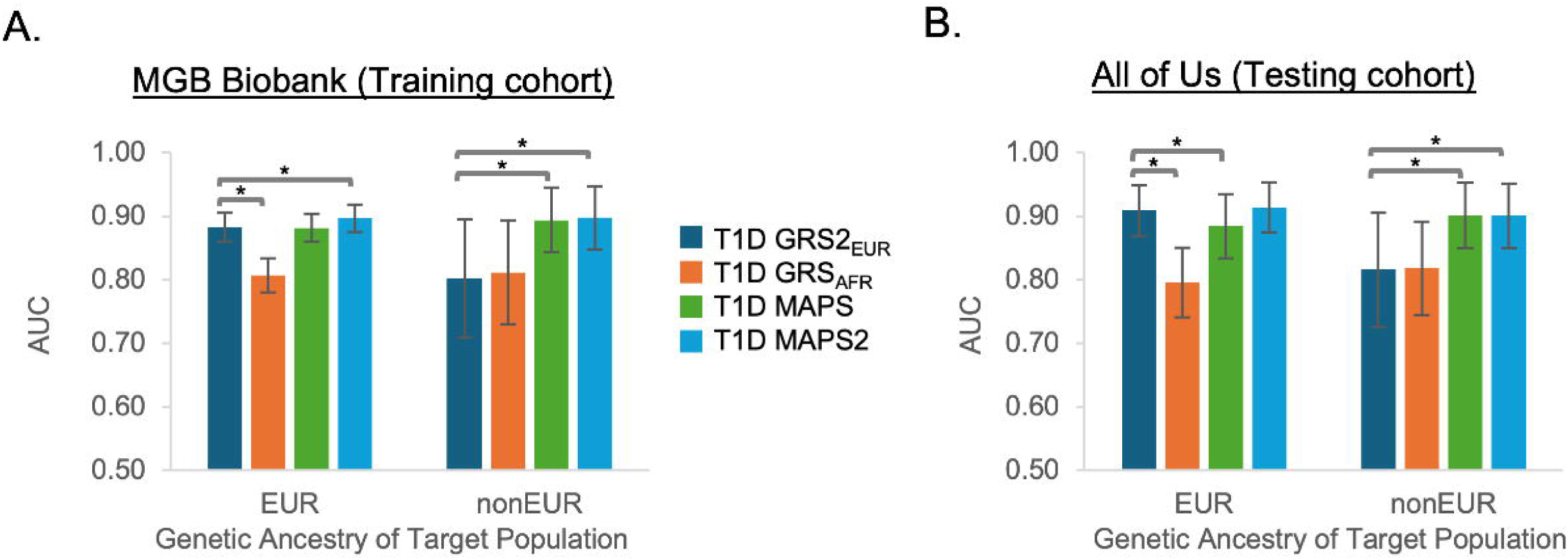
AUC of T1D polygenic scores. AUCs are displayed in (A) MGB Biobank (training cohort) and (B) All of Us (testing cohort). All AUCs were compared to the AUC of T1D GRS2_EUR_ using the DeLong test. Error bars denote 95% confidence interval. * *P* < 0.05.

### Validation of T1D MAPS

Next, to exclude the possibility of overfitting, we applied T1D MAPS in an external cohort. We validated the score using the All of Us Research Program, which contained over 100,000 individuals with genomic data, including 86 individuals with strictly defined type 1 diabetes (61 with EUR ancestry and 25 with non-EUR ancestry; Supplementary Table S1). We used whole genome sequencing data to assemble multi-locus HLA haplotypes. Once again, we found that the genetic variants used to tag HLA haplotypes in T1D GRS2_EUR_ were highly correlated with sequencing-based haplotypes among individuals with EUR ancestry, but the correlation was weaker in non-EUR ancestry (mean R^2^ 0.92 vs. 0.78, Wilcoxon *P* = 1.6 x 10^−3^; Supplementary Fig. S1B).

In the testing cohort, T1D MAPS had slightly lower AUC compared to T1D GRS2_EUR_ within EUR ancestry (0.89 vs. 0.91, *P* = 0.02; Fig. 2B). However, T1D MAPS2 performed similarly to T1D GRS2_EUR_ within EUR ancestry (0.91 vs. 0.91, *P* = 0.45). Among non-EUR ancestry, T1D MAPS and T1D MAPS2 were each superior to T1D GRS2_EUR_ (0.90 vs. 0.82, *P* = 0.04; 0.90 vs. 0.82, *P* = 0.04) and to T1D GRS_AFR_ (0.90 vs. 0.82, *P* = 7.3 x 10^−3^; 0.90 vs. 0.82, *P* = 7.1 x 10^−3^). When we analyzed the HLA and nonHLA components separately, we observed similar patterns in both the training and testing cohorts (Supplementary Fig. S2). In addition, after controlling for four principal components, our primary findings were unchanged (Supplementary Fig. S3). Finally, we confirmed that T1D MAPS could accurately distinguish between individuals with type 1 and type 2 diabetes (Supplementary Fig. S4).

Because the testing cohort had a low number of individuals with type 1 diabetes, we also tested a more lenient phenotype definition, which did not account for age at diabetes onset^30^. This lenient definition included 569 individuals with type 1 diabetes (351 with EUR ancestry and 218 with non-EUR ancestry). Using this definition, T1D MAPS and T1D GRS2_EUR_ performed similarly within EUR ancestry (AUC 0.82 vs. 0.83, *P* = 0.66). Interestingly, both scores had low performance within non-EUR ancestry, with no significant difference between T1D MAPS and T1D GRS2_EUR_ (AUC 0.67 vs. 0.67, *P* = 0.59), or between T1D MAPS and T1D GRS_AFR_ (AUC 0.67 vs. 0.67, *P* = 0.93).

### Determination of optimal score cutoffs

After developing T1D MAPS, we assessed the optimal score threshold for discriminating between individuals with and without type 1 diabetes. First, we confirmed that the distribution of each polygenic score may differ based on genetic ancestry (Fig. 3). For example, among individuals with type 1 diabetes, the mean T1D GRS2_EUR_ is lower in non-EUR ancestry compared to EUR ancestry. In contrast, both T1D MAPS and T1D MAPS2 had similar score distributions among individuals with type 1 diabetes, although there were slight ancestry-related differences among individuals without type 1 diabetes.

**Figure 3.**
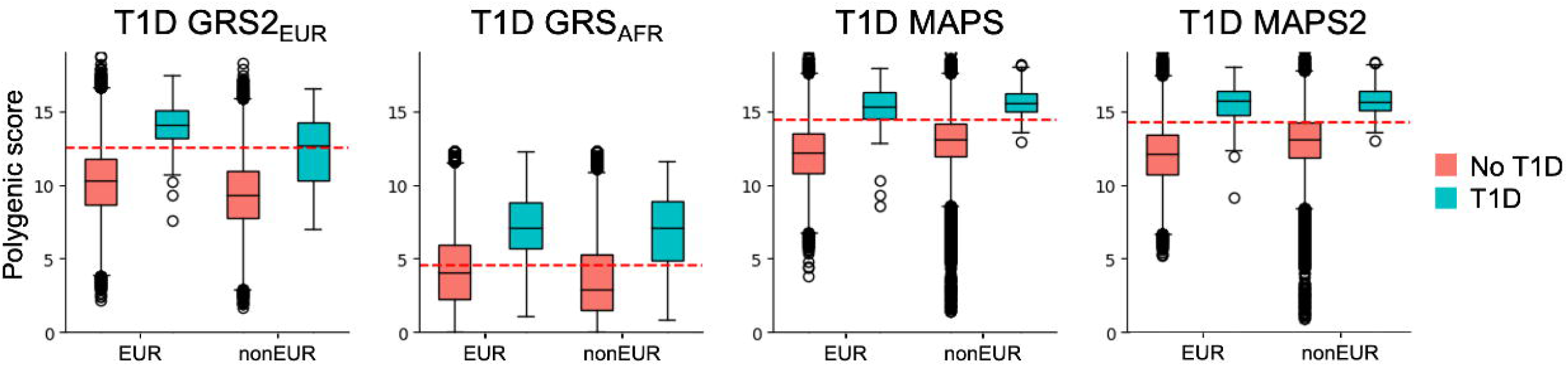
Distribution of T1D polygenic scores. Boxplots display the distribution of each polygenic score in All of Us, stratified by type 1 diabetes status and genetic ancestry. The dashed red line in each graph represents the score with the maximum Youden index.

To quantify the performance of each score, we calculated the sensitivity, specificity, and Youden index (sensitivity + specificity – 1) at various score thresholds in All of Us (Table 1). Once again, we demonstrated that score performance may differ based on genetic ancestry. For example, for T1D GRS2_EUR_, a score of 12.35 (85^th^ percentile in the overall population) yielded a Youden index of 0.72 in EUR ancestry but only 0.45 in non-EUR ancestry. In comparison, for T1D MAPS2, a score of 14.47 (85^th^ percentile in the overall population) yielded a Youden index of 0.69 in EUR ancestry and 0.67 in non-EUR ancestry.

**Table 1.**
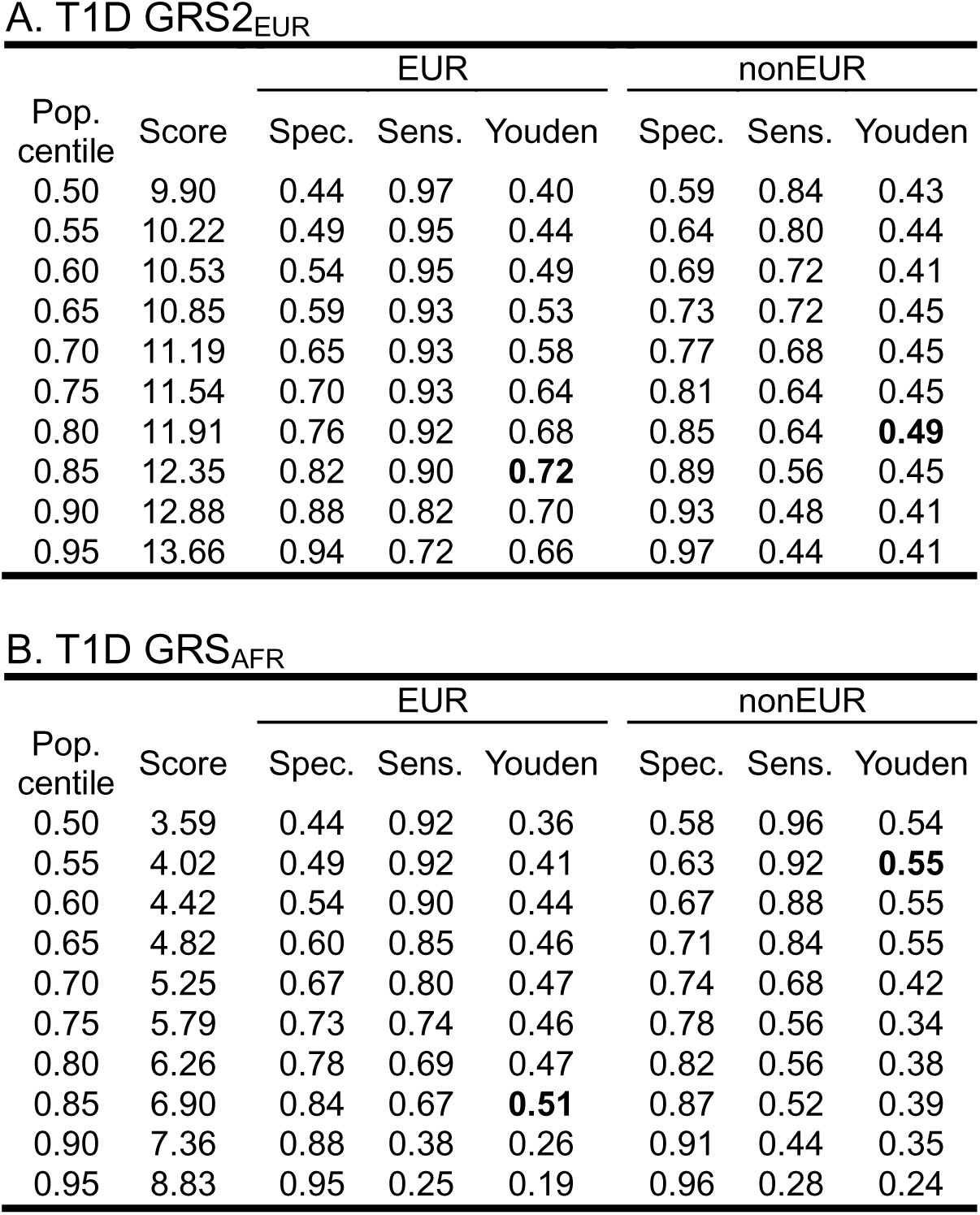
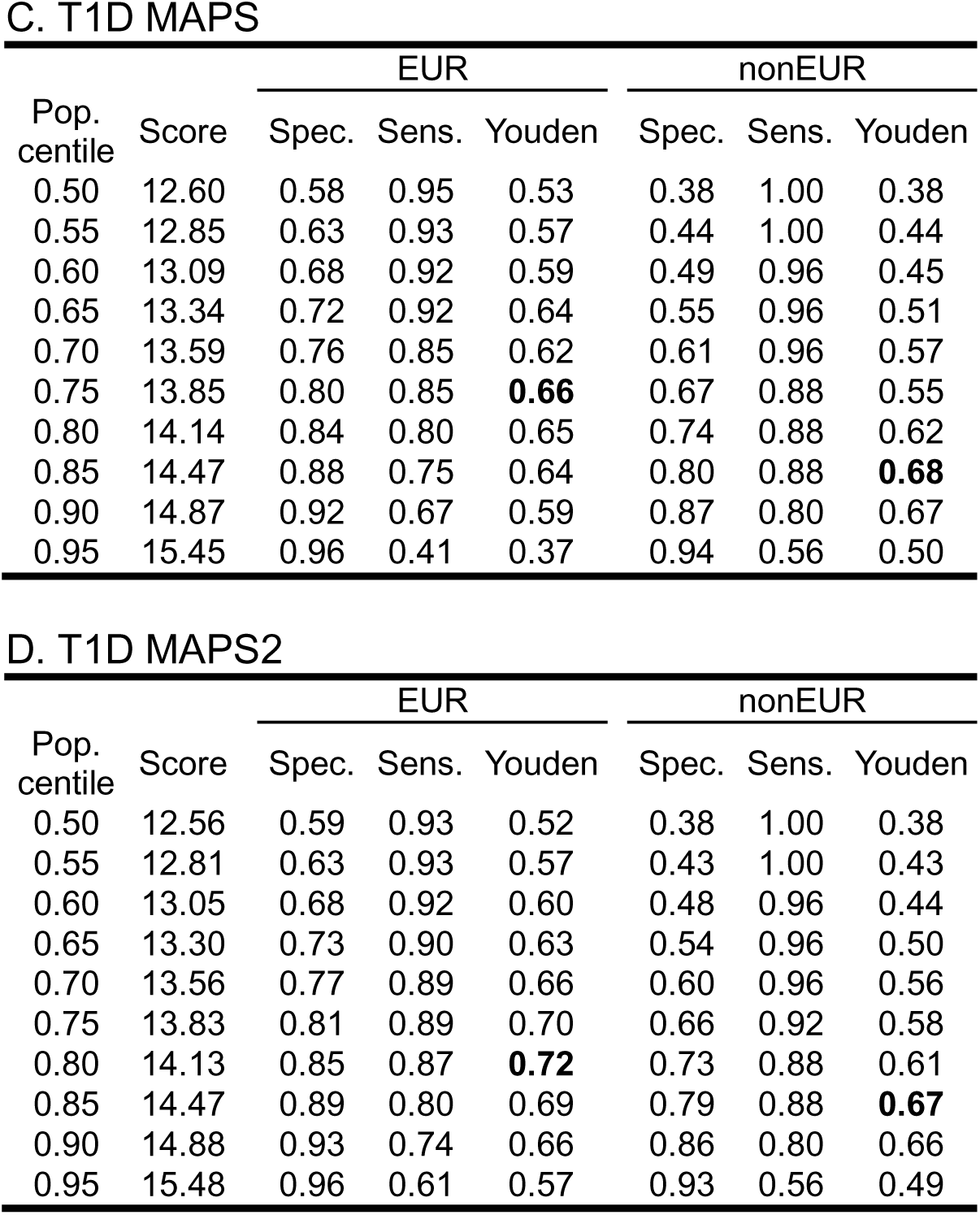
Sensitivity and specificity of T1D polygenic scores. This table displays the sensitivity, specificity, and Youden index (sensitivity + specificity – 1) for various values of each polygenic score, as applied in the testing cohort (All of Us). The centile for each score was calculated for the entire population and was then applied across all ancestry groups. The maximum Youden index for each column is denoted in bold font.

## Conclusions

Polygenic scores are highly predictive of type 1 diabetes risk. Here, we demonstrated that a novel multi-ancestry polygenic score can improve type 1 diabetes prediction in individuals with non-EUR genetic ancestry, while maintaining high predictive power in EUR ancestry populations.

Our results highlight the critical necessity of accurately capturing HLA haplotypes. In particular, we demonstrated that the genetic variants used to tag HLA haplotypes in T1D GRS2_EUR_ are more closely correlated with HLA haplotypes in EUR compared to non-EUR ancestry. We used two alternative methods to determine HLA haplotypes: imputation from genotype array data and assembly from whole genome sequencing data. Each resulting HLA score was highly correlated with type 1 diabetes risk, confirming previous findings that approaches using genotype arrays or whole genome sequencing have similar predictive power for type 1 diabetes^12^. Additionally, we demonstrated how a global extended polygenic score can improve disease prediction by incorporating numerous genetic variants that do not achieve genome-wide significance in a GWAS.

Although T1D MAPS demonstrated strong predictive power across ancestry groups, the AUC was marginally lower than T1D GRS2_EUR_ in individuals with EUR ancestry in the testing cohort. Therefore, we developed an alternative score, T1D MAPS2, which also accounts for an individual’s predicted proportion of EUR genetic ancestry. This approach avoids the drawbacks of existing polygenic scores because it can be applied in all settings and does not require individuals to declare their self-reported race or ethnicity. Furthermore, T1D MAPS2 models genetic ancestry as a continuous variable, avoiding the need to sort individuals into discrete categories such as EUR or AFR.

In practice, clinicians may use polygenic scores to identify individuals at high risk of developing type 1 diabetes, or to distinguish between type 1 and type 2 diabetes for individuals with atypical presentations^37^. In both circumstances, it is useful to designate a score cutoff to separate high-risk and low-risk individuals. However, identifying the optimal cutoff is challenging, as the underlying distribution of each polygenic score may vary based on genetic ancestry^12^. One option is to introduce ancestry-specific score thresholds; nevertheless, even with tailored thresholds, predictive power may differ among ancestry groups, and this approach requires classification of individuals into discrete categories. Here, we demonstrate that a multi-ancestry score allows for a single threshold with high discriminatory power across all ancestry groups.

Although our results are promising, our findings were limited by small sample size. In particular, we examined a relatively low number of individuals with type 1 diabetes and non-EUR ancestry in both the training and testing cohorts. This limited our ability to detect differences between T1D MAPS and other polygenic scores; nevertheless, despite our limited statistical power, we observed significantly higher performance with T1D MAPS in non-EUR ancestry, demonstrating the value of this score. Future studies should replicate these findings in larger cohorts. In addition, our analysis of non-EUR ancestry groups was limited to individuals with AFR and AMR ancestry. Future studies should include individuals from other ancestral backgrounds, such as East Asian or South Asian.

Our assessment of disease risk relies on accurate classification of type 1 diabetes. One option is use to focus on individuals with positive autoantibodies; however, most individuals in our cohorts did not have autoantibody testing, and in any case up to 15% of individuals with type 1 diabetes do not have detectable autoantibodies^38^. Instead, we applied classification algorithms using electronic medical records, but such algorithms are less accurate in non-EUR ancestry, compared to EUR ancestry^28^. In MGB Biobank, we mitigated this issue by manually reviewing clinical notes, but clinical notes were not available in All of Us. Interestingly, when we analyzed a more lenient classification algorithm in All of Us, we found that all polygenic scores (including T1D GRS2_EUR_ and T1D MAPS) had poor discriminatory power within non-EUR ancestry. This highlights challenges associated with identifying type 1 diabetes in diverse settings, particularly because diabetes may present differently across global populations^39^.

From a practical perspective, implementing T1D MAPS may require more resources compared to T1D GRS2_EUR_ or T1D GRS_AFR_^40^. In particular, imputation or assembly of HLA haplotypes requires an extra step beyond standard processing of genomic data. In addition, calculating a global extended polygenic score with one million variants requires more computational resources compared to a limited score with fewer than one hundred variants. Nevertheless, the variants required to compute T1D MAPS can be obtained by performing imputation with the TOPMED reference panel^25^ on data from commercially available genotype arrays, which are becoming increasingly affordable.

Similarly, T1D MAPS2 requires additional computational resources to determine predicted probabilities for each continental ancestry group. This approach aligns an individual’s genetic data to external reference panels^31^ and does not vary based on the underlying genetic admixture of a given population. However, if continuous probabilities of genetic ancestry are not readily available, then T1D MAPS can be used instead, with relatively similar performance.

Overall, we demonstrated that a multi-ancestry polygenic score can accurately predict type 1 diabetes risk across ancestry groups, with a single score threshold that identifies high-risk individuals. This score is useful in clinical practice and may advance knowledge of diabetes risk, particularly in underrepresented populations. Future work should replicate these findings in larger cohorts and expand to include individuals from other ancestry groups.

## Data Availability

Data from the All of Us Research Program are available to authorized users on the All of Us Researcher Workbench. Data from MGB Biobank are only available to MGB-affiliated researchers with approval from the MGB Institutional Review Board. The weights files and code used to generate T1D MAPS and T1D MAPS2 are available at https://github.com/snam-mgh/T1D_MAPS. The code used to generate T1D GRS2_EUR is available at https://github.com/sethsh7/PRSedm/. The weights used to calculate T1D GRS_AFR are available in the Polygenic Score Catalog (https://www.pgscatalog.org) under score ID PGS000023.

## Personal Thanks

We gratefully acknowledge All of Us participants for their contributions, without whom this research would not have been possible. We also thank the National Institutes of Health’s All of Us Research Program for making available the participant data examined in this study.

## Funding

AJD is supported by National Institutes of Health (NIH) / National Institute of Diabetes and Digestive and Kidney Diseases (NIDDK) K23 DK140643. RJK is supported by NIH/NIDDK T32 DK007699. AKM, JMM, and MSU are supported by NIH / National Human Genome Research Institute (NHGRI) U01HG011723. JMM is supported by American Diabetes Association grant #11-22-ICTSPM-16, by the NIH / NIDDK under Award Number R01DK137993 and U01 DK140757, AMP CMD award from RFP 6 from the Foundation for the National Institutes of Health, and a Medical University of Bialystok (MUB) grant from the Ministry of Science and Higher Education (Poland). This work is supported by the Novo Nordisk Foundation (NNF21SA0072102). MSU is supported by Doris Duke Foundation Award 2022063, NIDDK U54DK118612, and NIDDK U01DK140757.

## Conflicts of Interest

MSU is involved in a research collaboration with Novo Nordisk that is unrelated to content of this manuscript.

## Author Contributions

AJD, JCF, JMM, and MSU designed the study. AJD performed data analysis and wrote the initial draft of the manuscript. ASB, DAM, and SN contributed to data analysis. ABB, RN, YL, and AAM-R assisted with generating HLA haplotypes. AH-C and RM assisted with determining genetic ancestry. DAM, SO-G, and SSR contributed to study design and generated association statistics in the Type 1 Diabetes Genetics Consortium. RJK, SAS, RAO, and AKM contributed to discussion and reviewed and edited the manuscript. All authors approved the final version of the manuscript. MSU is the guarantor of this work and, as such, had full access to all the data in the study and takes responsibility for the integrity of the data and the accuracy of the data analysis.

## Prior Presentation

Parts of this study were presented in abstract form at the 85^th^ Scientific Sessions of the American Diabetes Association, Chicago, IL, 20-23 June 2025.

## Data and Resource Availability

Data from the All of Us Research Program are available to authorized users on the All of Us Researcher Workbench. Data from MGB Biobank are only available to MGB-affiliated researchers with approval from the MGB Institutional Review Board. The weights files and code used to generate T1D MAPS and T1D MAPS2 are available at https://github.com/snam-mgh/T1D_MAPS. The code used to generate T1D GRS2_EUR_ is available at https://github.com/sethsh7/PRSedm/. The weights used to calculate T1D GRS_AFR_ are available in the Polygenic Score Catalog (https://www.pgscatalog.org) under score ID PGS000023.

## Notes

### Author Declarations

Analysis of Mass General Brigham Biobank was approved by the Mass General Brigham IRB (study protocol 2016P001018). Analysis of the All of Us cohort was approved by an institutional Data Use and Registration Agreement between Mass General Brigham and the All of Us Research Program (study protocol 2020P002213).

