## Supplementary Figures for "Development and Validation of a Type 1 Diabetes Multi-Ancestry Polygenic Score"

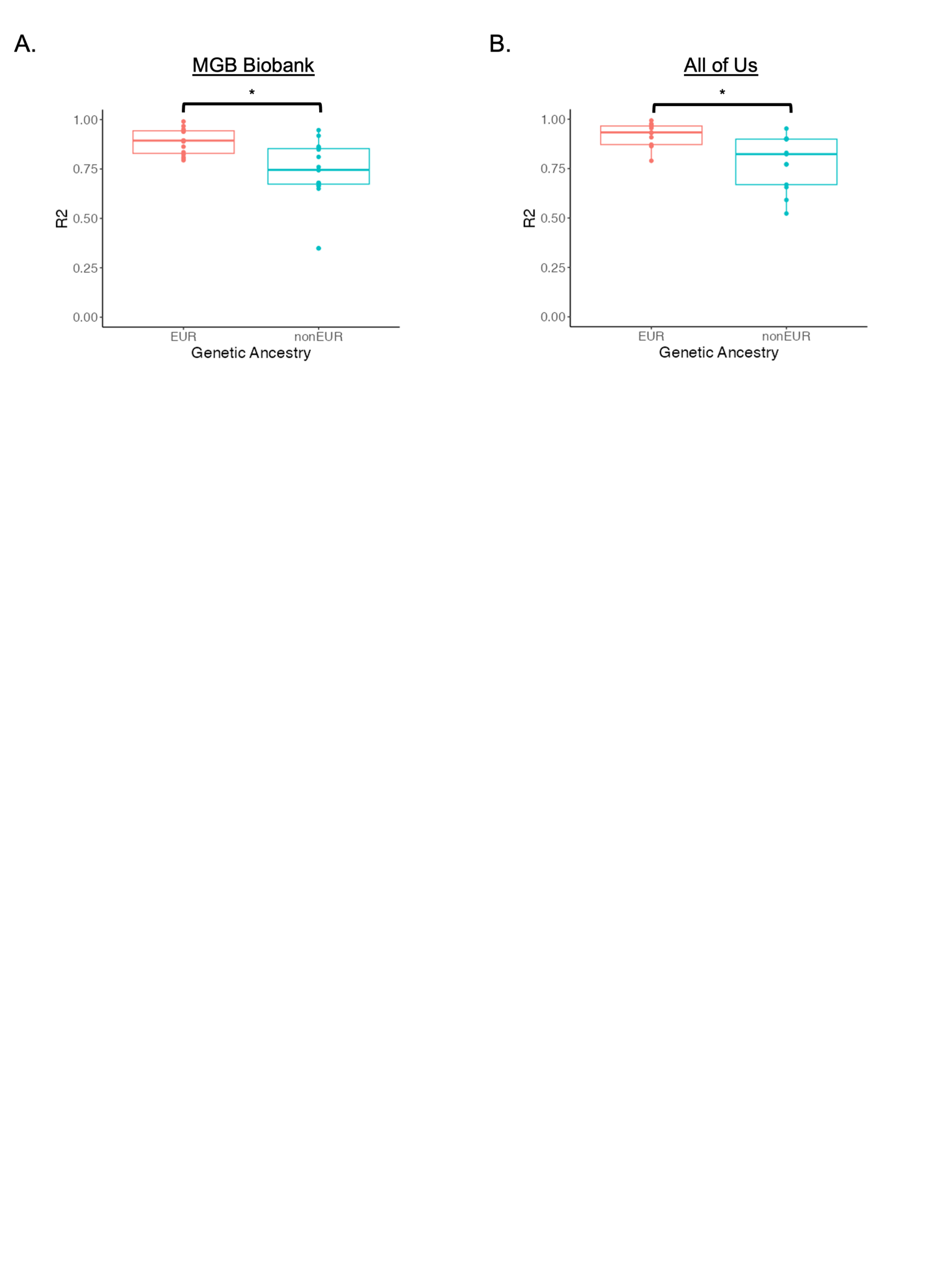

**Supplementary Figure S1. Correlation between tag variants and HLA haplotypes**

We generated HLA haplotypes using (A) imputation in MGB Biobank or (B) whole genome sequencing in All of Us. In both cohorts, we also assigned HLA haplotypes based on 14 tag variants used in T1D GRS2_EUR_. Each boxplot shows the square of the Pearson correlation coefficient (R^2^), demonstrating the correlation between haplotypes generated by tag variants and those generated by imputation or whole genome sequencing. * *P* < 0.05.

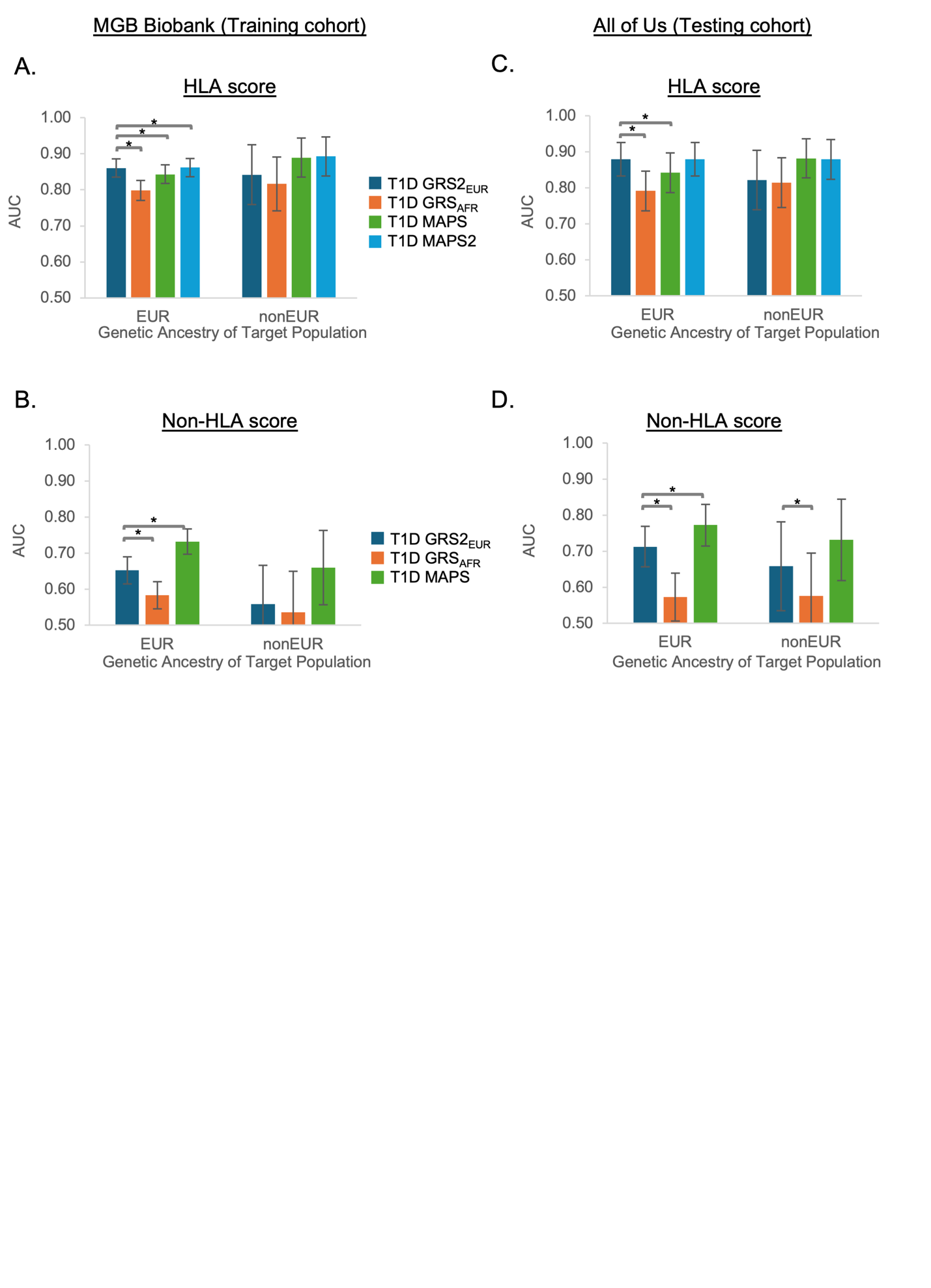

**Supplementary Figure S2. AUC of T1D polygenic score components**

AUCs are displayed separately for HLA and non-HLA components of each polygenic score in (A,B) MGB Biobank (training cohort) and (C,D) All of Us (testing cohort). All AUCs were compared to the AUC of T1D GRS2_EUR_ using Delong’s test. Error bars denote 95% confidence interval. * *P* < 0.05.

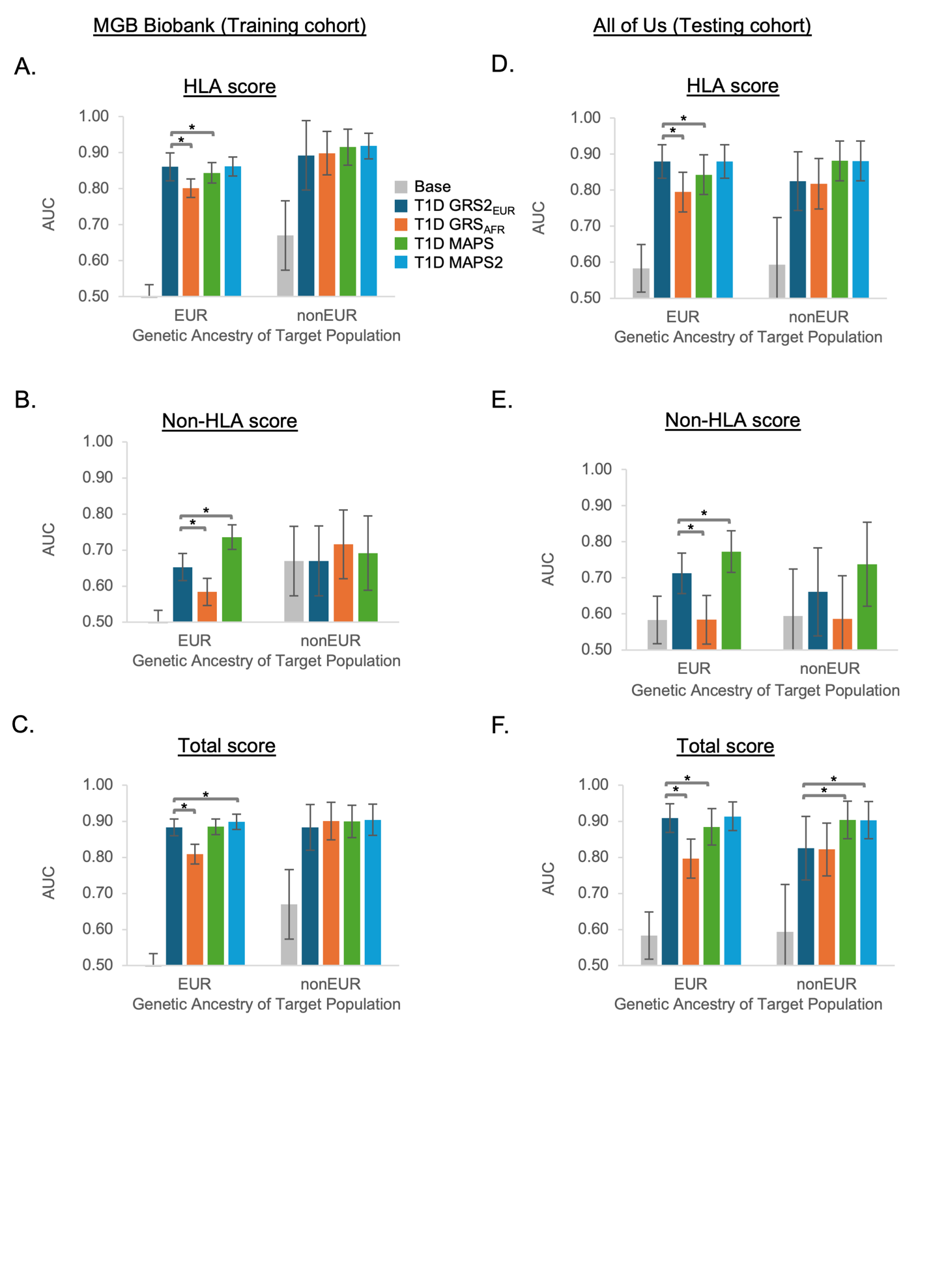
**Supplementary Figure S3. AUC of T1D polygenic scores, controlling for genetic ancestry**

We calculated the AUC of each polygenic score after controlling for four principal components of genetic ancestry. AUCs are displayed separately for the HLA component, non-HLA component, or total score in (A-C) MGB Biobank (training cohort) and (D-F) All of Us (testing cohort). The base model refers to a logistic regression model with covariates only (no polygenic score). All AUCs were compared to the AUC of T1D GRS2_EUR_ using Delong’s test. Error bars denote 95% confidence interval. * *P* < 0.05.

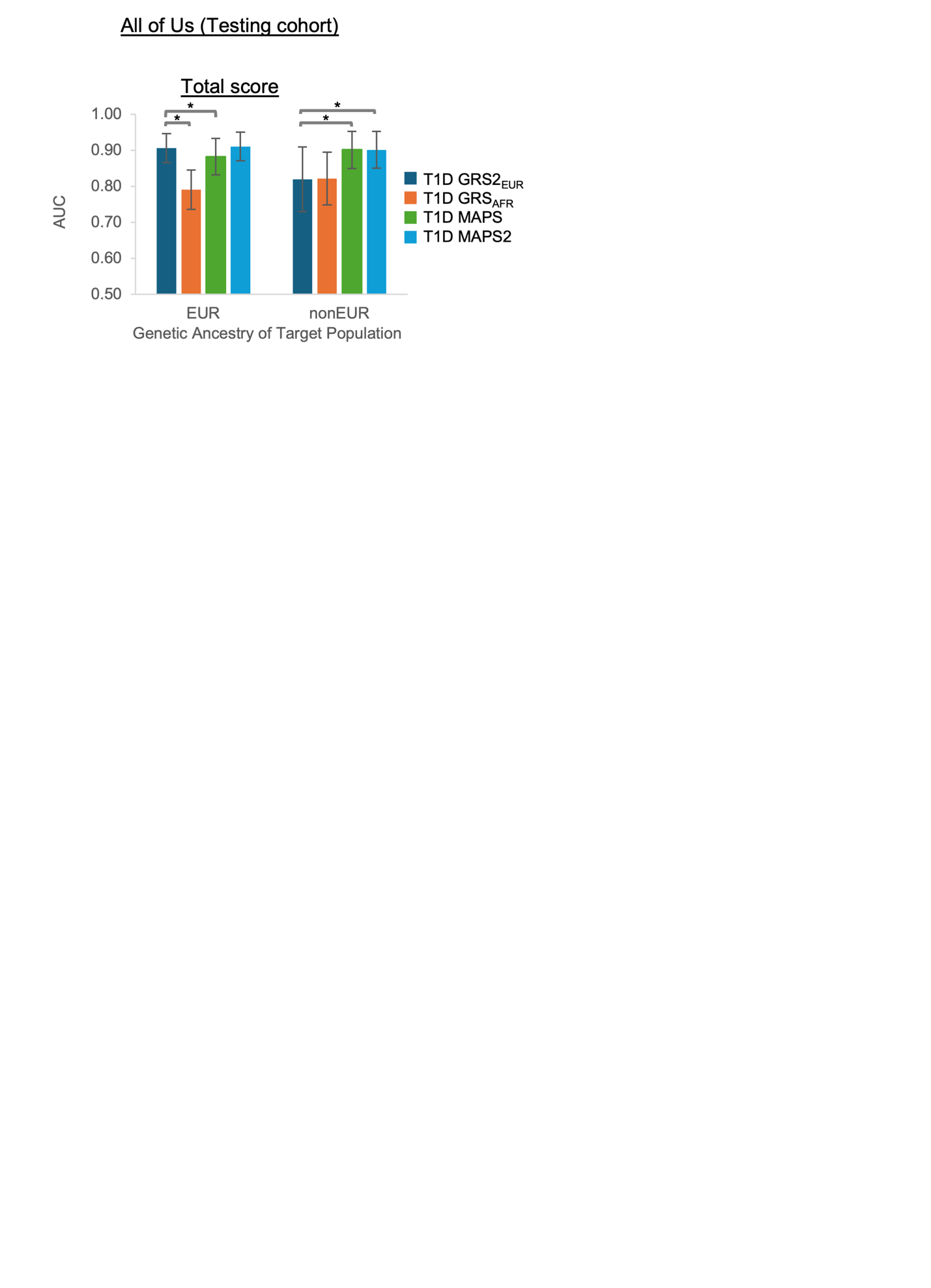

**Supplementary Figure S4. Ability of T1D polygenic scores to discriminate between type 1 and type 2 diabetes**

We calculated the AUC of each polygenic score to discriminate between individuals with type 1 or type 2 diabetes in All of Us (testing cohort). All AUCs were compared to the AUC of T1D GRS2_EUR_ using Delong’s test. Error bars denote 95% confidence interval. * *P* < 0.05.
